# Analysis and perception of Instagram posts referring to diastasis recti abdominis, exercise and sports-related content: a mixed-method study protocol

**DOI:** 10.1101/2023.07.05.23292242

**Authors:** S Giagio, T Rial Rebullido, S Salvioli, T Innocenti, P Pillastrini, IS Moore, GM Donnelly

## Abstract

**Objectives:** This study will evaluate the health information posted in Instagram (IG) concerning in diastasis recti abdominis (DRA), exercise and sports, and it will explore how IG content is perceived and impacts the daily lives of women with DRA.

**Methods:** A convergent parallel mixed methods design will be used. Through an observational study, public IG posts content written in English language will be descriptively analysed and reviewed using the QUality Evaluation Scoring Tool (QUEST) criteria. We will extract data on exercise and sport suggestions for women with DRA, along with any proposed indications and contraindications.

Eligible women with a diagnosis of DRA will be recruited via a worldwide e-survey using Qualtrics, and invited to take part to one-to-one interviews via Microsoft Teams. Semi-structured interviews will aim to explore their IG use and their perceptions on information related to DRA, exercise, and sports. Descriptive and thematic analysis will be conducted.

**Implications:** The availability and quality of information on social media are becoming increasingly important aspects of patient care. For sports medicine and women’s health clinicians, the findings of the present study will give an overview of DRA content that are easily accessible to women, underscoring the related posts characteristics. Furthermore, results could impact clinical practice by providing clinicians with a better understanding of patients’ perceptions and beliefs from IG DRA content, with the ultimate aim of better engaging with them and dispelling any potential misconceptions.

## INTRODUCTION

The use of the internet and social media can reduce the distance between patients and medical professionals, as well as offer a number of opportunities and advantages [1,2]. One of the most popular social media platforms for sharing and searching for biomedical information is Instagram (IG) [3,4]. Its user base is rapidly growing and disseminating content that can be accessed by anyone without the supervision and validation of experts or professionals. Unfiltered content can be used as a supposedly reliable source of health-related information [2,3], but there is a chance that non-evidence based and false claims will spread. Additionally, false and poor-quality content may persuade followers to engage in harmful or unhealthy behaviour [5].

Diastasis recti abdominis (DRA) is defined as the presence of divergence between the rectus abdominis muscles along the linea alba [6–8]. This condition is common in women [6,9], with a prevalence rate of 39 to 45.5% six months after childbirth and 32.6 % at one year [8]. Despite DRA’s negative impact on quality of life and body image [10], Gluppe et al. recently found that the majority of women learned about DRA by searching medical information and advice through social media [11]. Moreover, only a few women opted to contact a healthcare professional [11].

Evaluating the quality of health information in Instagram concerning DRA, exercise, and sports represents a significant health issue. This is due to the prevalence rates of DRA and the UK chief medical officers (CMOs) recommendation to stay active during and after childbirth [12], as well as the shift in pregnancy-related information seeking from caregivers to social media [11].

Furthermore, the impact and influence of these contents on the lives of women with DRA are still poorly understood and under researched.

Therefore, the primary aims of this mixed method study will be: 1) to evaluate the health information and suggestions in IG posts concerning DRA, exercise and sports through an observational study design; 2) to explore how women with DRA perceive IG content and to understand the impact of this content in their daily lives through a qualitative study design. The secondary objective will be to descriptively characterize IG creators and type of content.

## MATERIALS AND METHODS

### Study design

This study will adopt a convergent parallel mixed-methods design [13]. We will collect both quantitative and qualitative data in parallel, analyse them separately and compare results subsequently. For this reason, we will further report methods for each study design.

Reported information for each section will be reported as “Part 1: Social media posts about DRA” and “Part 2: Experiences of women with DRA”. See **Figure 1**.

**FIGURE 1.**
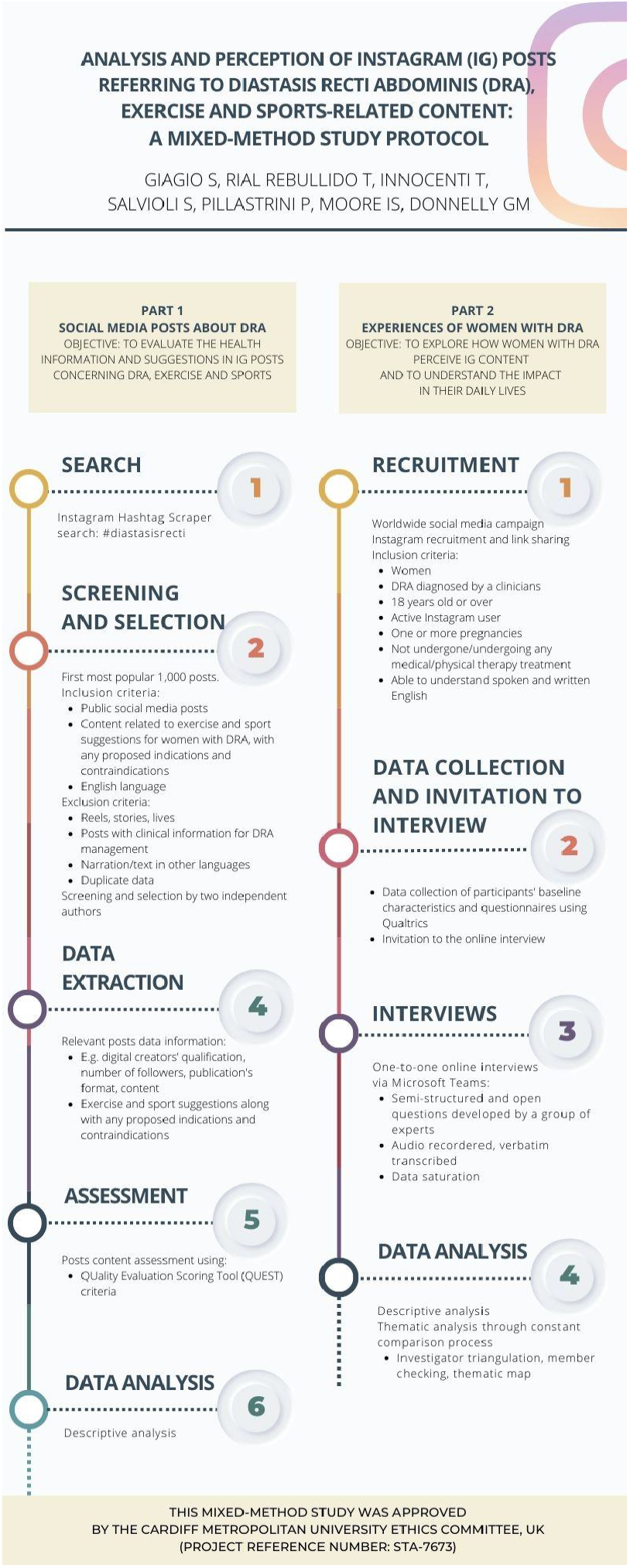
Flow-diagram: summary of the quantitative and qualitative process.

To ensure appropriateness of reporting, the Mixed Methods Article Reporting Standards (MMARS) will be used [14]. The protocol of this study was approved by the Cardiff Metropolitan University Ethics Committee, UK (Project Reference Number: Sta-7673; 30/06/2023).

### PART 1: SOCIAL MEDIA POSTS ABOUT DRA

An observational study will be conducted using a convenience sample of posts written in English language regarding DRA, exercise and sports-related content on Instagram via the online free Instagram Hashtag Scraper website.

#### Objectives

1. To evaluate the health information and suggestions in Instagram posts concerning DRA, exercise and sports.
2. Descriptively characterize IG creators and type of content.

#### Post selection criteria

##### Inclusion criteria

Only public social media posts with content related to exercise and sport suggestions for women with DRA, along with any proposed indications and contraindications, written in English language will be eligible for inclusion. To define exercise and sports, we use the terminology provided by the World Health Organization [15].

##### Exclusion criteria

Posts will be excluded if they contain registered lives, stories, reels or information not relevant to the objectives, narration/text in other languages, and duplicate data containing the same content. Given the aim of this study is to collect and analyse data on DRA sport and exercise suggestions (e.g., what could be indicated to practice or not), posts containing information and indications of clinical DRA management will not be considered. In addition, comments on the posts will not be extracted and analysed.

##### Process

Through Instagram Hashtag Scraper website, a comprehensive search for “#diastasisrecti” will be conducted independently by two authors (SG, SS). The search will be performed in parallel on a convenient day for both authors between 01 and 04 June 2023. The first 1.000 most popular posts will be independently screened for eligibility by the same two authors with any potential disagreement resolved by a third author (IM). The use of a single sensitive hashtag (#diastasisrecti) not including terminology related to exercise and sports is chosen based on a) the heterogeneity of hashtag uses in the social media platform related to women’s health; b) a preliminary social media search which showed the most common findings in accordance with the topic. Thereby, for eligibility, exercise and sports-related content will be evaluated manually.

To identify possible issues with collection and to ensure that there were no differences between the two searches, a pilot search was carried out on March, 31th 2023. **Figure 1** shows the entire process of screening and selection of posts.

#### Data extraction

Data will be extracted from the main accounts which posted the eligible posts. For example, related data will include the digital creators’ qualification, number of followers and main profile information will be collected. In particular:

a. Publication’s format: e.g. (a) text only (slide in the format of the picture containing only written text); (b) picture-text (single or multiple); (c) video-text; (d) others.
b. Content: exercise and sports suggestions for women with DRA, along with any indications and contraindications.

#### IG posts assessment

The quality of content will be assessed using the QUality Evaluation Scoring Tool (QUEST) criteria [16]. QUEST is a recent tool to evaluate online health information composed of a short set of criteria that can be used by health care professionals and researchers. Scores in the individual sections are weighted and summed to generate a total score of up to 28.

#### Statistical analysis

Data will be analysed using Statistical Package for the Social Sciences Inc. (SPSS software Chicago, IL, USA) to produce descriptive statistics. All findings will be presented as aggregated data.

### PART 2: EXPERIENCES OF WOMEN WITH DRA

A qualitative study design is suggested to explore the significance of numerous social variables that people encounter in their context [17]. This study design may be used to not only describe the various aspects of a phenomenon but also to propose unique viewpoints.

All procedures will adhere to the 1964 Helsinki Declaration and its later amendments or comparable ethical standards, as well as the ethical requirements of the institutional and/or national research committee. All participants will provide written informed consent and data protection policy before the study takes place. The consent can be withdrawn at any time. Participants will not be compensated for participation in this study.

#### Objective

- To explore how women with DRA perceive IG content and to understand the impact in their daily lives, through a qualitative study design.

### Participant and recruitment

As recommended by qualitative methodology researchers, we will use the principles of purposeful sampling, which consider for the identification and selection of information-rich cases, or people who have particular expertise in or experience with the phenomenon under investigation [18,19]. As a result, these individuals could help the researchers to maximize ef?cacy and validity [20].

#### Inclusion criteria

Women with DRA diagnosed by a clinician, age 18 years old or over, active Instagram user, who have had one or more pregnancies, who have not undergone or undergoing at the time of this study any medical/physical therapy treatment and are able to understand spoken and written English.

#### Exclusion criteria

Women who do not match the above inclusion criteria.

### Process, data collection and interviews

In July 2023, a social media campaign will be launched on the first author’s IG account (SG; silviagi.physio). A specific content (e.g. figure) will be created including a secure anonymous Qualtrics link to access the survey. We will expect that the content will be further disseminated and shared by other accounts.

Using Qualtrics, participants will be required to 1) confirm eligibility criteria, 2) complete the consent form prior to participating in the study, and 3) fill in basic demographic data and complete “The Diastasis Rectus Abdominis Questionnaire”. Participants will be then invited to schedule and take part in the interview. One-to-one interviews will be conducted via Microsoft Teams by GD. Interviews will last approximately 30 minutes and will be conducted using semi-structured and open questions developed by a group of experts (SG, IM, TR, GD).

The general aim of the interview will be to investigate personal experiences and the impact of IG content about DRA, exercise, and sports in daily life and sports participation. Additional questions will be asked concerning examples of information and suggestions (e.g., indications and contraindications for exercise and sports practice) that are recalled by participants. We will then explore further reasons and motivations behind their choice to follow or not the content creators’ suggestions or advice.

No a-priori sample size calculation is expected for qualitative design; interviews will be concluded when data saturation is reached.

Qualitative data collected via interview discussions will be recorded via Microsoft Teams. Any recordings will be stored on One Drive and password protected, verbatim transcribed (GD) and then deleted. At the point of transcription (i.e., before analysis) identifiable data will be de-identified using codenames to maintain confidentiality and data protection. Codenames are index terms assigned to participants, specifically we will use simple alphanumerical form, such as “ID1, ID2, etc”. Following this, thematic content analysis using Braun & Clarke’s [21] will be performed.

### Thematic analysis

Analysis will be conducted through constant comparison process [17]. The participant interviews will be coded by two independent authors (coders: SG, SS) to the point where no new categories will be discovered after the transcript analysis [22]. As reported above, data collection will end when the data saturation point is reached [23]. In order to identify themes, we will then proceed on to the analysis of the categories. Coders will then discuss findings and reach an agreement on a final thematic map. Finally, results will be discussed with all authors to obtain different perspectives on the data [24]. To achieve credibility, we will employ peer debriefing to provide a peer examination of the research process. Using member checking, participants will be given a chance to review the data collected by interviewers and the data’s interpretations. We will provide thick descriptions to ensure transferability. To demonstrate dependability, we will ensure that the research process is logical, traceable and clearly documented and we will use the code-recode procedure. To achieve conformability, investigator triangulation will be adopted. Each author from a different discipline (physical therapy, women’s health field, sports medicine, research methodology) will contribute to the discussion and understanding of the complex phenomenon under investigation [20,25].

## ETHICS AND DISSEMINATION

Approval from the Cardiff Metropolitan University Ethics Committee has been obtained (Project Reference Number: Sta-7673).

All raw data will be stored on a Cardiff Metropolitan password protected computer (OneDrive), anonymised, and only be accessible by the research team. Specifically for the quantitative data, all raw e-survey data will be collected and stored using Qualtrics (CMet license), and the IP address tracking will be turned off. All information relating to the raw data will be securely stored on Cardiff Metropolitan OneDrive for 10 years in accordance with Cardiff Metropolitan University’s ethics guidelines and destroyed thereafter and managed in line with the Data Protection Act 2018. https://www.gov.uk/government/collections/data-protection-act-2018.

For Part 1: We will present aggregated data and a summary of the content without reporting digital creators’ names, accounts, web URLs or any other identifier. We will comply to current *“Instagram Terms and Conditions”* protecting the privacy of Instagram users and creators.

Specifically for Part 2, all participants will be required to provide informed consent after being informed of the purpose and procedures of the study, risks and benefits, the right to withdraw, confidentiality and privacy regulations.

For interviews, all audio records will be deleted following analysis. All recordings will be stored on One-Drive (CMet license) and password protected. All transcripts will be anonymised prior to analysis using alphanumerical form (e.g. ID01 etc). A best effort to remove/anonymised identifiable information that could be disclosed during interview process will be made.

A manuscript will be prepared and submitted for publication in an appropriate peer-reviewed journal upon completion. The study findings will be disseminated at a relevant (inter)national conference. We believe that the results of this investigation will be relevant to clinicians in the field of sports medicine and women’s health.

## Data Availability

All data produced in the present work will be contained in the final manuscript.

## Acknowledgements

Not applicable.

